# Monitoring Report: Viral Gastroenteritis - August 2024 Data

**DOI:** 10.1101/2024.05.21.24307632

**Authors:** Samuel Gratzl, Brianna M Goodwin Cartwright, Patricia J Rodriguez, Charlotte Baker, Duy Do, Nicholas L Stucky

## Abstract

**Background:** Few sources regularly monitor hospitalizations associated with viral gastroenteritis. This study provides current hospitalization trends associated with two common viruses causing gastroenteritis: norovirus and rotavirus.

**Objective:** This study aims to supplement the surveillance data provided by the CDC by describing latest trends (through August 31, 2024) overall and for each virus. This study also provides valuable insight into two at-risk populations: infants and children (age 0-4) and older adults (age 65 and over).

**Methods:** Using a subset of real-world electronic health record (EHR) data from Truveta, a growing collective of health systems that provide more than 18% of all daily clinical care in the US, we identified people who were hospitalized between October 01, 2019 and August 31, 2024. We identified people who tested positive for each virus within 14 days of the hospitalization. We report monthly trends in the rate of hospitalizations associated with each virus per all hospitalizations and test positivity for the overall population and the two high-risk sub populations: infants and children and older adults.

**Results:** We included 8,153 hospitalizations of 7,939 unique patients who tested positive for a monitored virus between October 01, 2019 and August 31, 2024. Additionally, we included 354,235 lab results across 172,810 unique patients of which 15,741 were positive between October 01, 2019 and August 31, 2024.

Hospitalizations associated with these norovirus and rotavirus accounted for 0.04% of all hospitalizations in August 2024 (-18.2% from July 2024). The combined test positivity rate of these two enteric viruses was 3.6% in August 2024. For the population between age 0-4 years old, the hospitalizations associated with these two enteric viruses accounted for 0.08% of all hospitalizations in August (+196.7% from July 2024). The enteric virus test positivity rate was 10.3% for the month of August in this age group. In the population over 65 years of age, hospitalizations associated with these enteric viruses accounted for 0.04% of all hospitalizations in August 2024 (-26.0% from July 2024). The enteric virus test positivity rate was 2.4% for the month of August in this age group.

**Discussion:** We see evidence that we are in the nadir of the norovirus and rotavirus season for the overall population. The overall timeseries trends of hospitalizations associated with these viruses show a steady decrease over the last months, leading to a rate of 0.04% in August 2024 in the overall population. However, we see first indicators that the next season is already beginning for the population age 0-4 years old with an increase of the hospitalization rate of +196.7% from July.

In the overall and adult over age 65 population, norovirus causes 4 to 5 times higher hospitalization rates than rotavirus. However, in the population of infants and children, both viruses have similar hospitalization rates.

We will continue to monitor trends in common viruses causing gastroenteritis.

**Trends in Surveillance:** *Overall population:* 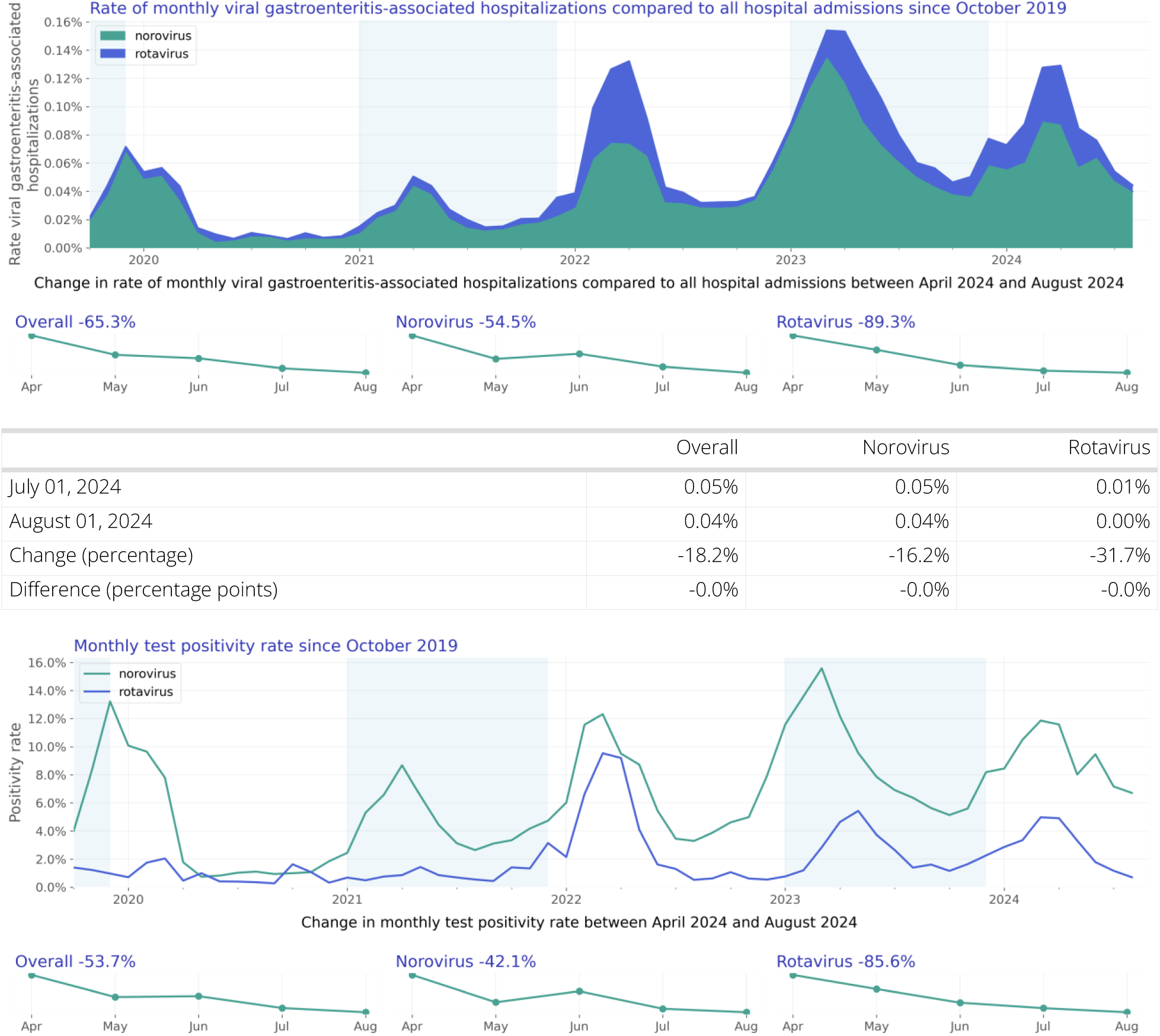

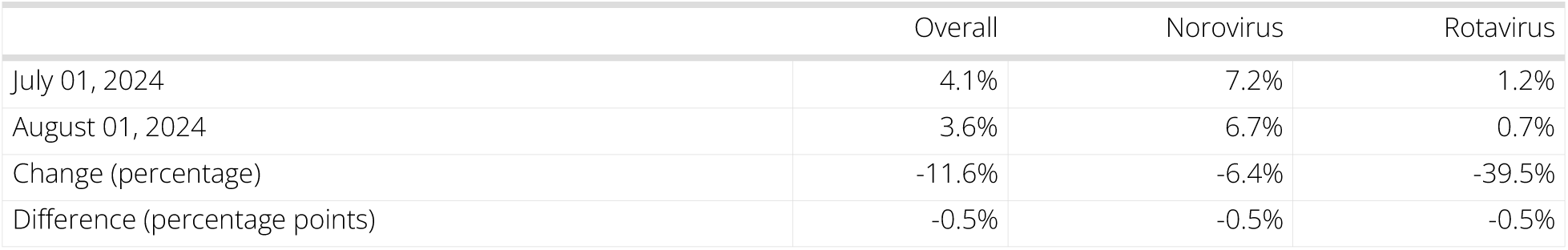 Overall, the rate of hospitalizations associated with these two enteric viruses decreased from July to August (-18.2%). Norovirus-related hospitalizations decreased less than rotavirus-related hospitalizations (-16.2% vs -31.7%). Combined hospitalizations associated with these enteric viruses accounted for 0.04% of all hospitalizations in August 2024. The combined test positivity rate of these two enteric viruses decreased from July to August to 3.6% (-11.6%). Norovirus test positivity rate decreased by 6.4%, while rotavirus test positivity rate decreased significantly by 39.5%.

*Infants and children (age 0-4):* 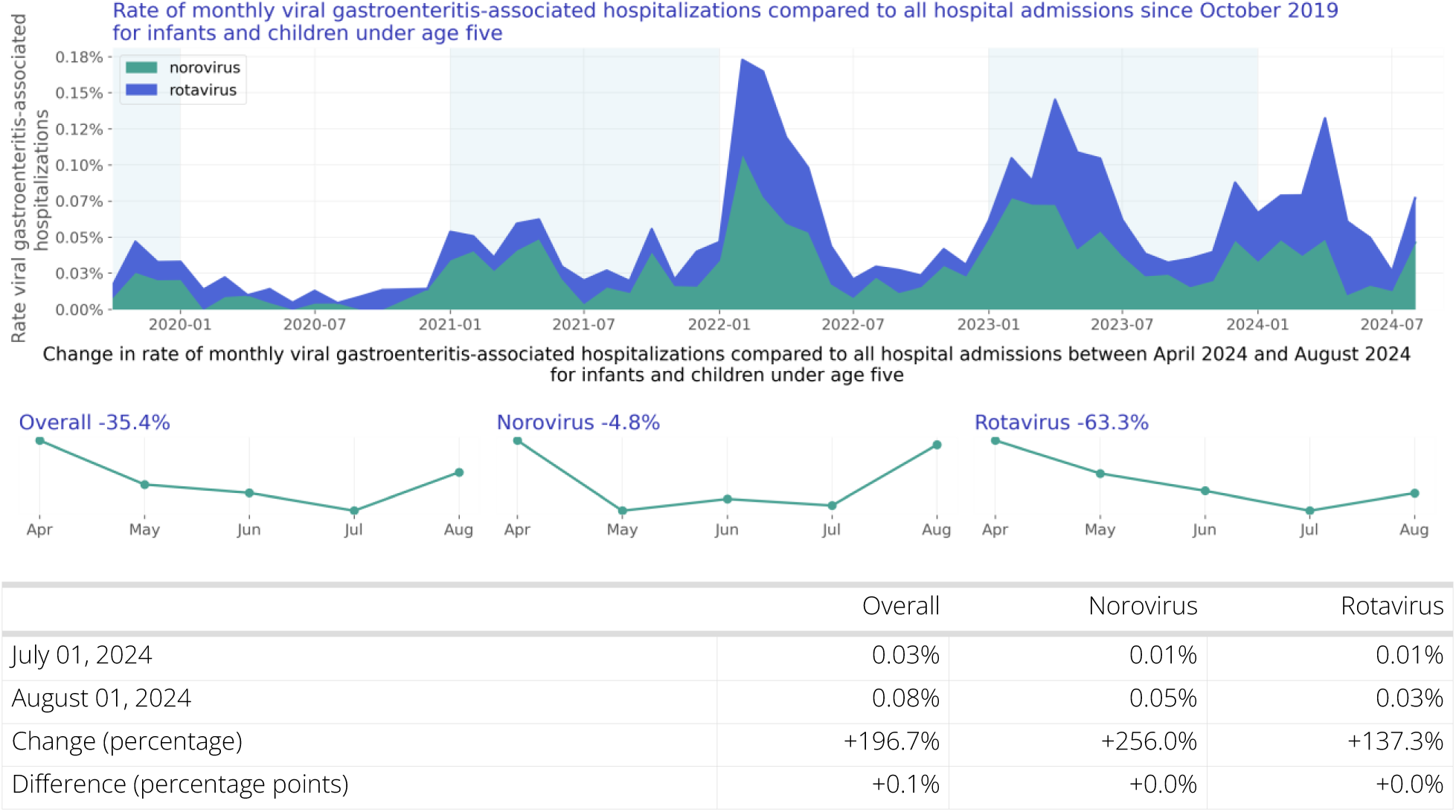

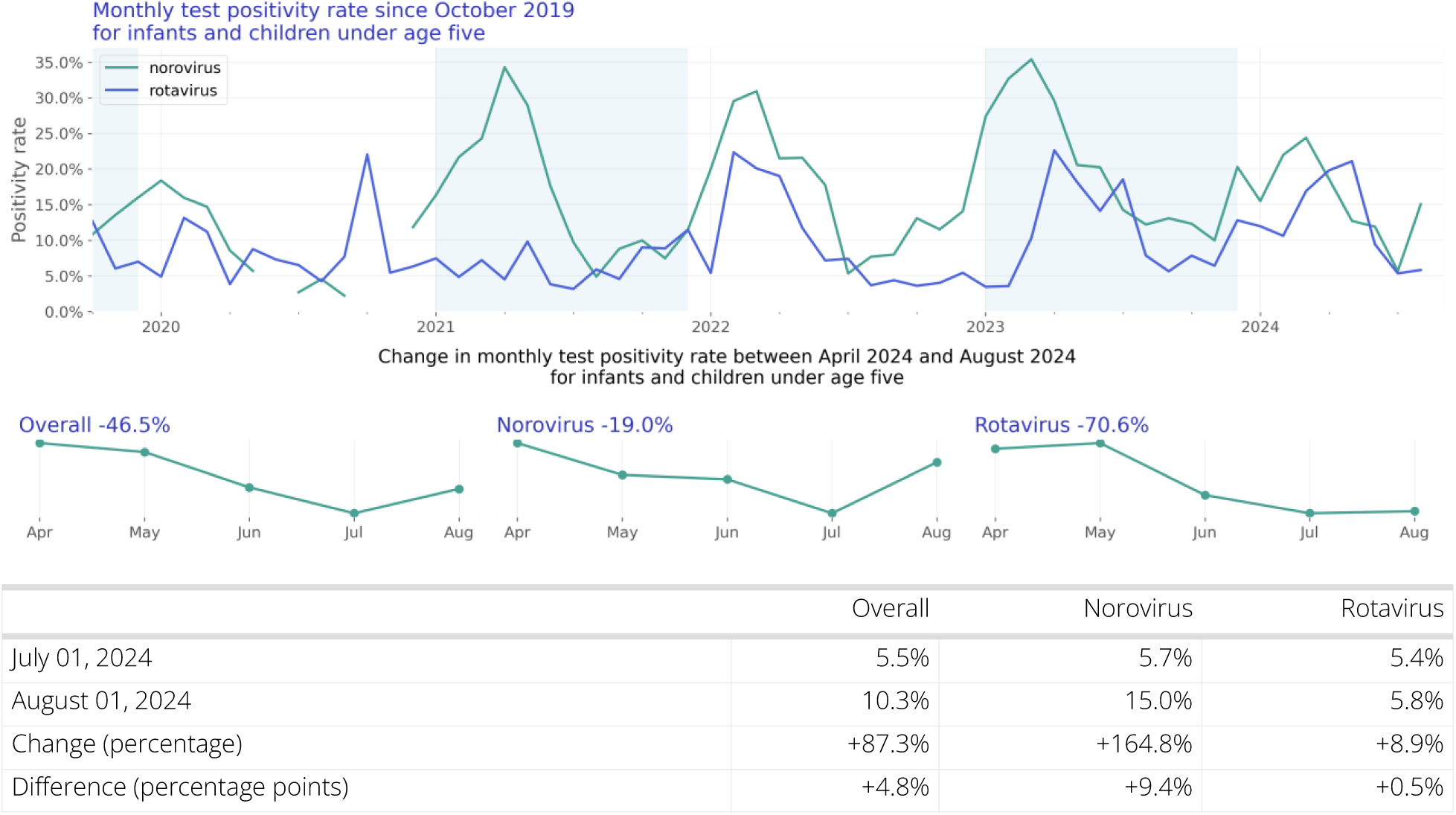 For the population between age 0-4 years old, the rate of hospitalizations associated with these two enteric viruses increased significantly from July to August (+196.7%). Both norovirus- and rotavirus-related hospitalizations increased (+256.0% and +137.3% respectively). Combined hospitalizations associated with these enteric viruses accounted for 0.08% of all hospitalizations in August in this age group. For this population, the enteric virus test positivity rate increased significantly from July to August (+87.3% change). Norovirus test positivity rate increased significantly by 164.8%, while rotavirus test positivity rate increased by 8.9%. The enteric virus test positivity rate was 10.3% for the month of August in this age group.

*Older adults (age 65 and over):* 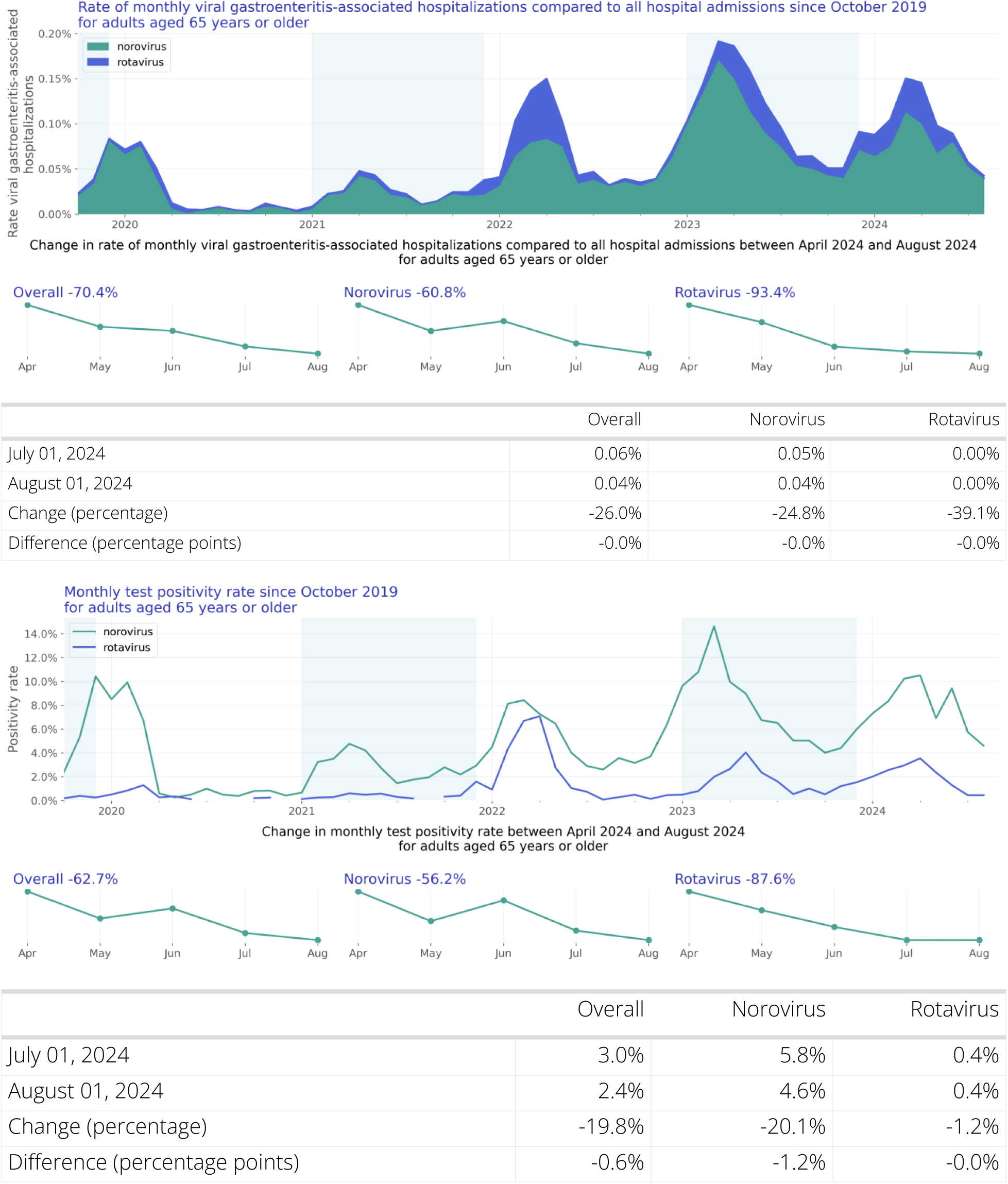 In the population over 65 years of age, there was a 26.0% decrease in the rate of hospitalizations associated with these two enteric viruses from July to August. Norovirus-related hospitalizations decreased (-24.8%), so did rotavirus-related hospitalizations (-39.1%). Combined hospitalizations associated with these enteric viruses accounted for 0.04% of all hospitalizations in August in this age group. For this population, the enteric virus test positivity rate decreased from July to August (- 19.8% change). Norovirus test positivity rate decreased by 20.1%, and rotavirus test positivity rate by 1.2%. The enteric virus test positivity rate was 2.4% for the month of August in this age group.

## Introduction

Norovirus and rotavirus are significant contributors to gastrointestinal infections worldwide, imposing substantial burdens on populations and healthcare systems (Troeger 2018, Ahmed 2014, Lopman 2011). In the United States, norovirus and rotavirus infections collectively account for a considerable proportion of hospitalizations related to gastrointestinal illnesses (Lopman 2011), underscoring the importance of comprehensively understanding the trends in viral infections affecting public health. Like their respiratory counterparts, norovirus and rotavirus infections pose particular risks to vulnerable populations, including infants, children, and older adults (Ahmed et al., 2014). Despite the focus on pediatric populations due to the severity of symptoms and potential complications, older adults also face significant risks of hospitalization and mortality from these viral infections (Anderson 2004). Severe outcomes and prolonged illness durations are particularly prevalent among the elderly. For hospitalized individuals who are immunocompromised or suffer from significant medical comorbidities, enteric virus infections can directly contribute to extended hospital stays, heightened medical complexities, and, in rare instances, mortality. These details highlight the interaction between viral infections and underlying health conditions.

It is important for public health experts and clinical providers to understand the trends in these infections to inform decisions about public health, clinical care, and public policy. Connecting population-level trends with granular clinical information available in Truveta Studio can be very useful to more deeply understand which cohorts are most impacted. This report is intended to supplement the surveillance data provided by the CDC (Centers for Disease Control and Prevention, 2023). This report includes additional independent data and clinical detail that is not captured in other reports.

## Methods

### Data

We evaluated viral gastroenteritis-associated hospitalization trends of two common viruses causing gastroenteritis - norovirus and rotavirus - between October 01, 2019 and August 31, 2024 using a subset of Truveta Data. Truveta provides access to continuously updated, and de-identified electronic health record (EHR) from a collective of US health care systems that provides 18% of all daily clinical care in the US, including structured information on demographics, encounters, diagnoses, vital signs (e.g., weight, BMI, blood pressure), medication requests (prescriptions), medication administration, laboratory and diagnostic tests and results (e.g., norovirus tests and values), and procedures. Updated EHR data are provided daily to Truveta by constituent health care systems. The data used in this study was provided on September 09, 2024 and included de-identified patient care data primarily located across ten states: Arizona, California, Florida, Illinois, Louisiana, Missouri, North Carolina, Texas, Virginia, and Washington.

### Population

We identified hospitalized patients who tested positive for one of the selected viruses causing gastroenteritis within 14 days before or during the hospitalization. Positive lab results were identified using LOINC codes. Every viral gastroenteritis-associated hospitalization has been grouped such that every hospitalization within 90 days is considered to be the same infection and thus only counted once.

In addition, we identified all lab results for one of the selected viruses and calculated the test positivity rate per month.

### Hospitalization rate analysis

Characteristics of viral gastroenteritis-associated hospitalized patients were summarized, including demographics and comorbidities by year. Characteristics are provided for the overall population, individual viruses, and two at-risk sub populations: infants and children (age 0-4) and older adults (age 65 and over).

Viral gastroenteritis-associated hospitalizations rates were summarized over time by virus. The hospitalization rate was calculated monthly as the number of patients with a viral gastroenteritis associated hospitalization divided by the number of patients with a hospital admission in that month. Patients were included in this calculation on the first day of their hospitalization. If their stay was greater than one day, they were not counted on subsequent dates.

Given the unadjusted nature of the data, the rates do not account for undertesting and other variability that exists across patient groups, providers, and systems. For further limitations, see the section below.

### Test positivity rate analysis

Lab results with known results were summarized over time by virus. The positivity rate was calculated monthly as the number of positive lab results divided by the total number of lab results with known results in that month.

Given the unadjusted nature of the data, the rates do not account for undertesting and other variability that exists across patient groups, providers, and systems. For further limitations, see the section below.

## Results

### Overall population

Our study population consists of 8,153 hospitalizations of 7,939 unique patients (Table 1).

**Table 1:** Patient characteristics by year.

|  | 2019 | 2020 | 2021 | 2022 | 2023 | 2024 | Overall |
| --- | --- | --- | --- | --- | --- | --- | --- |
| N | 319<br>(100.00%) | 510<br>(100.00%) | 729<br>(100.00%) | 1,845<br>(100.00%) | 2,873<br>(100.00%) | 1,663<br>(100.00%) | 7,939<br>(100.00%) |
| Age Group |  |  |  |  |  |  |  |
| < 6 months | 8 (2.51%) | 11 (2.16%) | 24 (3.29%) | 39 (2.11%) | 35 (1.22%) | 23 (1.38%) | 140 (1.76%) |
| 6 - <12 months | 3 (0.94%) | 6 (1.18%) | 32 (4.39%) | 30 (1.63%) | 43 (1.50%) | 28 (1.68%) | 142 (1.79%) |
| 1 - <2 years | 6 (1.88%) | 8 (1.57%) | 29 (3.98%) | 49 (2.66%) | 56 (1.95%) | 25 (1.50%) | 173 (2.18%) |
| 2 - 4 years | 6 (1.88%) | 7 (1.37%) | 18 (2.47%) | 54 (2.93%) | 74 (2.58%) | 35 (2.10%) | 194 (2.44%) |
| 5 - 17 years | 4 (1.25%) | 9 (1.76%) | 23 (3.16%) | 62 (3.36%) | 81 (2.82%) | 55 (3.31%) | 234 (2.95%) |
| 18 - 49 years | 71 (22.26%) | 111 (21.76%) | 171 (23.46%) | 375 (20.33%) | 556 (19.35%) | 347 (20.87%) | 1,631 (20.54%) |
| 50 - 64 years | 83 (26.02%) | 115 (22.55%) | 143 (19.62%) | 370 (20.05%) | 571 (19.87%) | 302 (18.16%) | 1,584 (19.95%) |
| 65 - 74 years | 51 (15.99%) | 81 (15.88%) | 109 (14.95%) | 380 (20.60%) | 592 (20.61%) | 322 (19.36%) | 1,535 (19.33%) |
| 75 - 84 years | 53 (16.61%) | 105 (20.59%) | 120 (16.46%) | 297 (16.10%) | 524 (18.24%) | 332 (19.96%) | 1,431 (18.02%) |
| 85+ years | 34 (10.66%) | 57 (11.18%) | 60 (8.23%) | 189 (10.24%) | 341 (11.87%) | 194 (11.67%) | 875 (11.02%) |
| Sex |  |  |  |  |  |  |  |
| Female | 163 (51.10%) | 266 (52.16%) | 385 (52.81%) | 973 (52.74%) | 1,505 (52.38%) | 901 (54.18%) | 4,193 (52.82%) |
| Male | 154 (48.28%) | 238 (46.67%) | 337 (46.23%) | 854 (46.29%) | 1,332 (46.36%) | 744 (44.74%) | 3,659 (46.09%) |
| Unknown | 2 (0.63%) | 6 (1.18%) | 7 (0.96%) | 18 (0.98%) | 36 (1.25%) | 18 (1.08%) | 87 (1.10%) |

| Race |  |  |  |  |  |  |  |
| --- | --- | --- | --- | --- | --- | --- | --- |
| White | 252<br>(79.00%) | 362<br>(70.98%) | 513<br>(70.37%) | 1,384<br>(75.01%) | 2,081<br>(72.43%) | 1,136<br>(68.31%) | 5,728<br>(72.15%) |
| Black or African American | 28 (8.78%) | 50 (9.80%) | 81<br>(11.11%) | 147 (7.97%) | 253 (8.81%) | 188<br>(11.30%) | 747 (9.41%) |
| Asian | 3 (0.94%) | 9 (1.76%) | 18 (2.47%) | 28 (1.52%) | 68 (2.37%) | 21 (1.26%) | 147 (1.85%) |
| Native Hawaiian or Other<br>Pacific Islander | 0 | 2 (0.39%) | 4 (0.55%) | 16 (0.87%) | 18 (0.63%) | 8 (0.48%) | 48 (0.60%) |
| American Indian or Alaska<br>Native | 0 | 5 (0.98%) | 6 (0.82%) | 19 (1.03%) | 25 (0.87%) | 14 (0.84%) | 69 (0.87%) |
| Other Race | 11 (3.45%) | 28 (5.49%) | 59 (8.09%) | 106 (5.75%) | 186 (6.47%) | 138 (8.30%) | 528 (6.65%) |
| Unknown | 25 (7.84%) | 54<br>(10.59%) | 48 (6.58%) | 145 (7.86%) | 242 (8.42%) | 158 (9.50%) | 672 (8.46%) |
| Ethnicity |  |  |  |  |  |  |  |
| Hispanic or Latino | 23 (7.21%) | 55<br>(10.78%) | 102<br>(13.99%) | 225<br>(12.20%) | 413<br>(14.38%) | 259<br>(15.57%) | 1,077<br>(13.57%) |
| Not Hispanic or Latino | 266<br>(83.39%) | 398<br>(78.04%) | 571<br>(78.33%) | 1,443<br>(78.21%) | 2,210<br>(76.92%) | 1,302<br>(78.29%) | 6,190<br>(77.97%) |
| Unknown | 30 (9.40%) | 57<br>(11.18%) | 56 (7.68%) | 177 (9.59%) | 250 (8.70%) | 102 (6.13%) | 672 (8.46%) |
| Virus |  |  |  |  |  |  |  |
| norovirus | 295<br>(92.48%) | 435<br>(85.29%) | 603<br>(82.72%) | 1,350<br>(73.17%) | 2,329<br>(81.07%) | 1,246<br>(74.92%) | 6,258<br>(78.83%) |
| rotavirus | 24 (7.52%) | 75<br>(14.71%) | 126<br>(17.28%) | 495<br>(26.83%) | 544<br>(18.93%) | 417<br>(25.08%) | 1,681<br>(21.17%) |

### Hospitalization rate over time

The rate of viral gastroenteritis-associated hospitalizations compared to all hospitalizations is shown in Figure 1. Figure 2 shows the same data stacked to represent the combined impact of the viruses.

**Figure 1:**
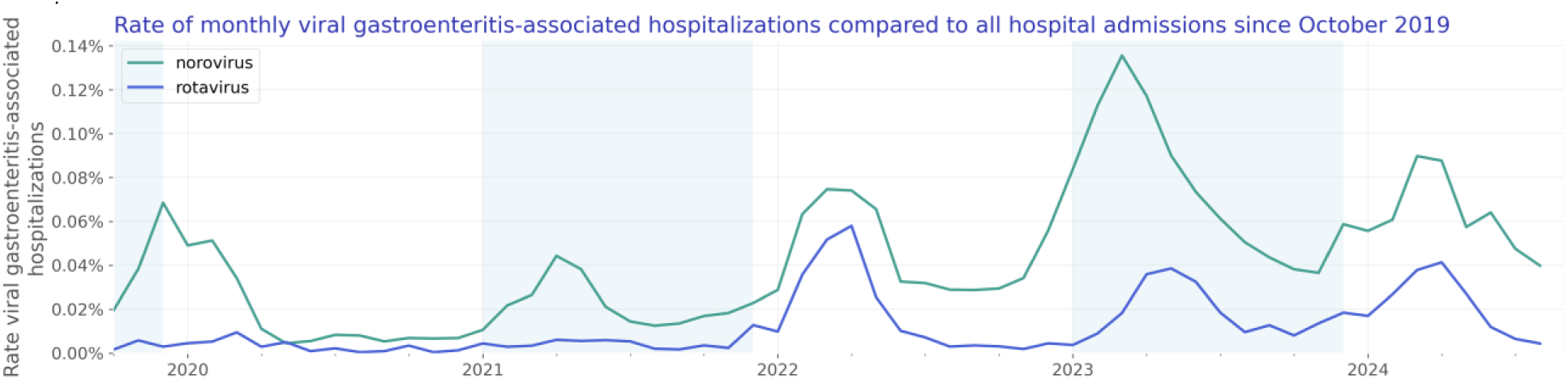
Rate of monthly viral gastroenteritis-associated hospitalizations compared to all hospital admissions since October 2019

**Figure 2:**
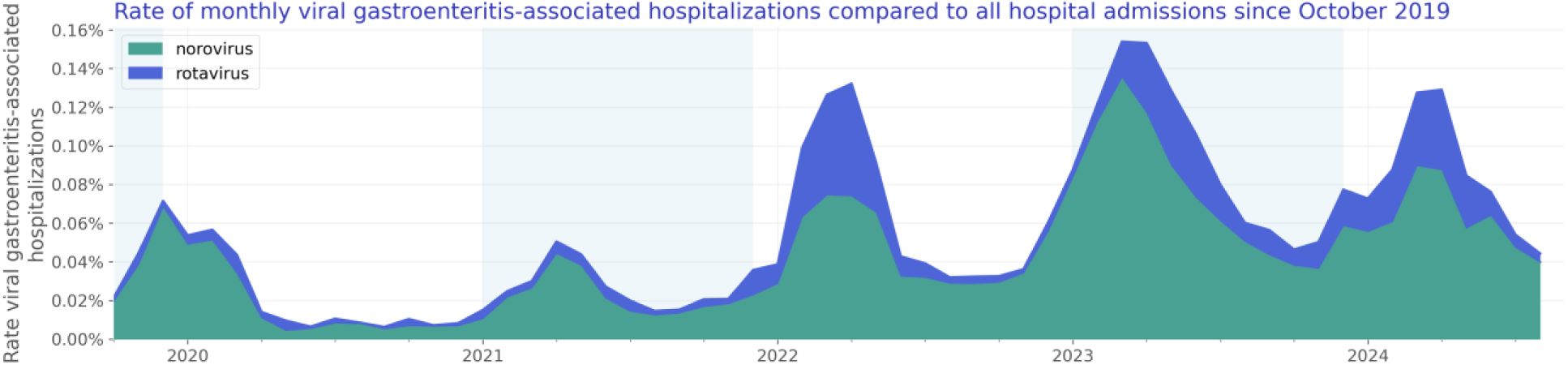
Rate of monthly viral gastroenteritis-associated hospitalizations compared to all hospital admissions since October 2019

### Test positivity rate over time

We included 354,235 lab results with known results of which 15,741 were positive. The test positivity rate is shown in Figure 3.

**Figure 3:**
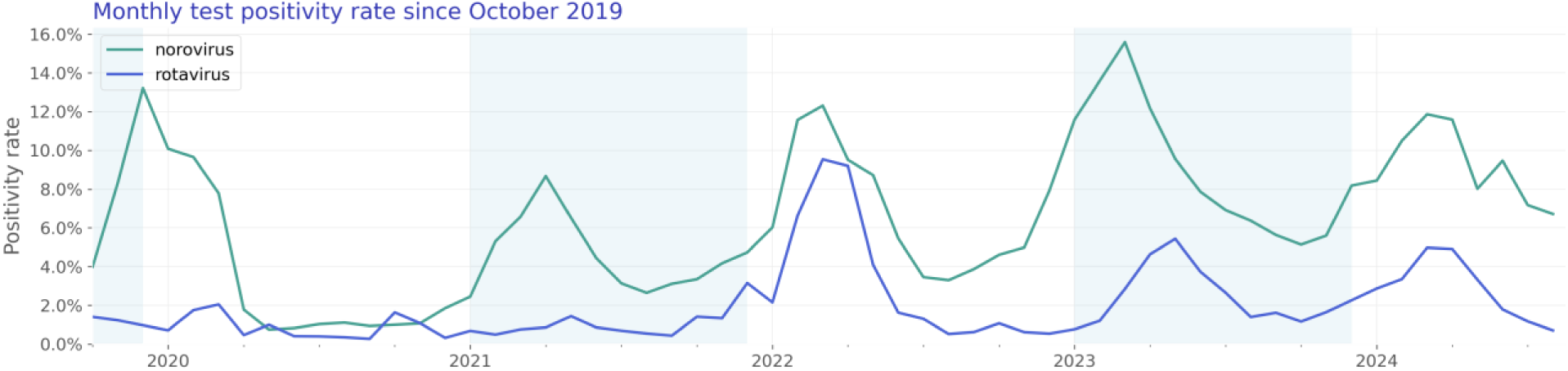
Monthly test positivity rate since October 2019

### Norovirus

Our norovirus study population consists of 6,368 hospitalizations of 6,258 unique patients (Table 2).

**Table 2:** Norovirus patient characteristics by year.

|  | 2019 | 2020 | 2021 | 2022 | 2023 | 2024 | Overall |
| --- | --- | --- | --- | --- | --- | --- | --- |
| N | 295<br>(100.00%) | 435<br>(100.00%) | 603<br>(100.00%) | 1,350<br>(100.00%) | 2,329<br>(100.00%) | 1,246<br>(100.00%) | 6,258<br>(100.00%) |
| Age Group |  |  |  |  |  |  |  |
| < 6 months | 4 (1.36%) | 3 (0.69%) | 12 (1.99%) | 24 (1.78%) | 14 (0.60%) | 6 (0.48%) | 63 (1.01%) |
| 6 - <12 months | 1 (0.34%) | 3 (0.69%) | 26 (4.31%) | 21 (1.56%) | 24 (1.03%) | 14 (1.12%) | 89 (1.42%) |
| 1 - <2 years | 4 (1.36%) | 3 (0.69%) | 23 (3.81%) | 29 (2.15%) | 39 (1.67%) | 17 (1.36%) | 115 (1.84%) |
| 2 - 4 years | 4 (1.36%) | 4 (0.92%) | 9 (1.49%) | 30 (2.22%) | 48 (2.06%) | 14 (1.12%) | 109 (1.74%) |
| 5 - 17 years | 4 (1.36%) | 7 (1.61%) | 13 (2.16%) | 35 (2.59%) | 53 (2.28%) | 35 (2.81%) | 147 (2.35%) |
| 18 - 49 years | 68 (23.05%) | 101 (23.22%) | 156 (25.87%) | 295 (21.85%) | 459 (19.71%) | 281 (22.55%) | 1,360 (21.73%) |
| 50 - 64 years | 80<br>(27.12%) | 101<br>(23.22%) | 120<br>(19.90%) | 281<br>(20.81%) | 474<br>(20.35%) | 227<br>(18.22%) | 1,283<br>(20.50%) |
| 65 - 74 years | 47<br>(15.93%) | 72<br>(16.55%) | 89<br>(14.76%) | 269<br>(19.93%) | 483<br>(20.74%) | 248<br>(19.90%) | 1,208<br>(19.30%) |
| 75 - 84 years | 52<br>(17.63%) | 92<br>(21.15%) | 103<br>(17.08%) | 226<br>(16.74%) | 442<br>(18.98%) | 255<br>(20.47%) | 1,170<br>(18.70%) |
| 85+ years | 31<br>(10.51%) | 49<br>(11.26%) | 52 (8.62%) | 140<br>(10.37%) | 293<br>(12.58%) | 149<br>(11.96%) | 714<br>(11.41%) |
| Sex |  |  |  |  |  |  |  |
| Female | 153<br>(51.86%) | 227<br>(52.18%) | 325<br>(53.90%) | 724<br>(53.63%) | 1,226<br>(52.64%) | 682<br>(54.74%) | 3,337<br>(53.32%) |
| Male | 140<br>(47.46%) | 202<br>(46.44%) | 273<br>(45.27%) | 613<br>(45.41%) | 1,071<br>(45.99%) | 549<br>(44.06%) | 2,848<br>(45.51%) |
| Unknown | 2 (0.68%) | 6 (1.38%) | 5 (0.83%) | 13 (0.96%) | 32 (1.37%) | 15 (1.20%) | 73 (1.17%) |
| Race |  |  |  |  |  |  |  |
| White | 235<br>(79.66%) | 315<br>(72.41%) | 426<br>(70.65%) | 1,017<br>(75.33%) | 1,717<br>(73.72%) | 868<br>(69.66%) | 4,578<br>(73.15%) |
| Black or African American | 24 (8.14%) | 47<br>(10.80%) | 74<br>(12.27%) | 110 (8.15%) | 203 (8.72%) | 141<br>(11.32%) | 599 (9.57%) |
| Asian | 3 (1.02%) | 5 (1.15%) | 14 (2.32%) | 18 (1.33%) | 54 (2.32%) | 13 (1.04%) | 107 (1.71%) |
| Native Hawaiian or Other<br>Pacific Islander | 0 | 2 (0.46%) | 3 (0.50%) | 11 (0.81%) | 8 (0.34%) | 4 (0.32%) | 28 (0.45%) |
| American Indian or Alaska<br>Native | 0 | 4 (0.92%) | 6 (1.00%) | 13 (0.96%) | 18 (0.77%) | 10 (0.80%) | 51 (0.81%) |
| Other Race | 10 (3.39%) | 18 (4.14%) | 40 (6.63%) | 74 (5.48%) | 129 (5.54%) | 92 (7.38%) | 363 (5.80%) |
| Unknown | 23 (7.80%) | 44<br>(10.11%) | 40 (6.63%) | 107 (7.93%) | 200 (8.59%) | 118<br>(9.47%) | 532 (8.50%) |
| Ethnicity |  |  |  |  |  |  |  |
| Hispanic or Latino | 18 (6.10%) | 38 (8.74%) | 69<br>(11.44%) | 150<br>(11.11%) | 295<br>(12.67%) | 169<br>(13.56%) | 739<br>(11.81%) |
| Not Hispanic or Latino | 249<br>(84.41%) | 350<br>(80.46%) | 487<br>(80.76%) | 1,072<br>(79.41%) | 1,826<br>(78.40%) | 995<br>(79.86%) | 4,979<br>(79.56%) |
| Unknown | 28 (9.49%) | 47<br>(10.80%) | 47 (7.79%) | 128 (9.48%) | 208 (8.93%) | 82 (6.58%) | 540 (8.63%) |

### Hospitalization rate over time

The rate of norovirus-associated hospitalization is shown in Figure 4. Figure 5 shows yearly trends.

**Figure 4:**
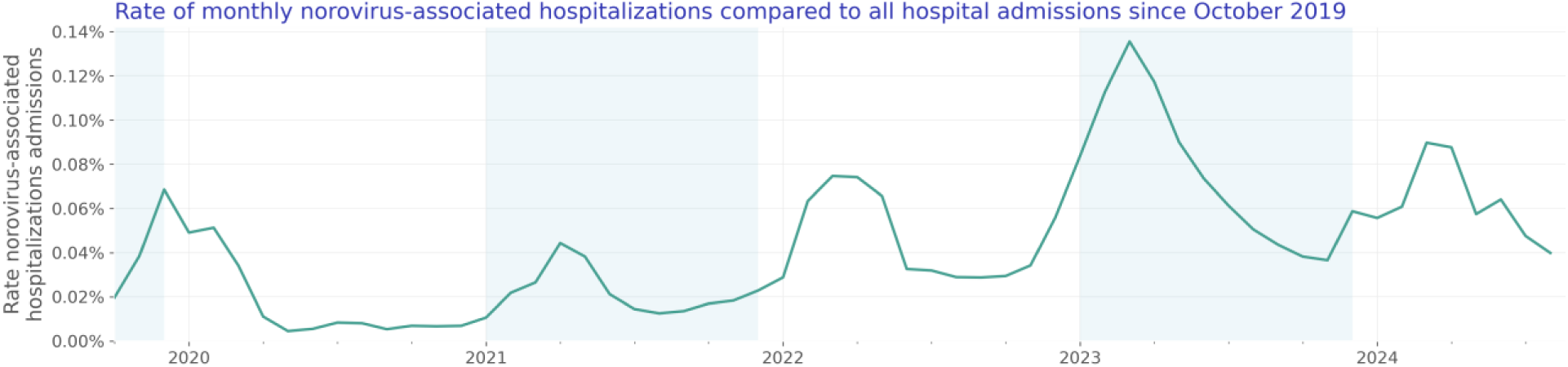
Rate of monthly norovirus-associated hospitalizations compared to all hospital admissions since October 2019

**Figure 6:**
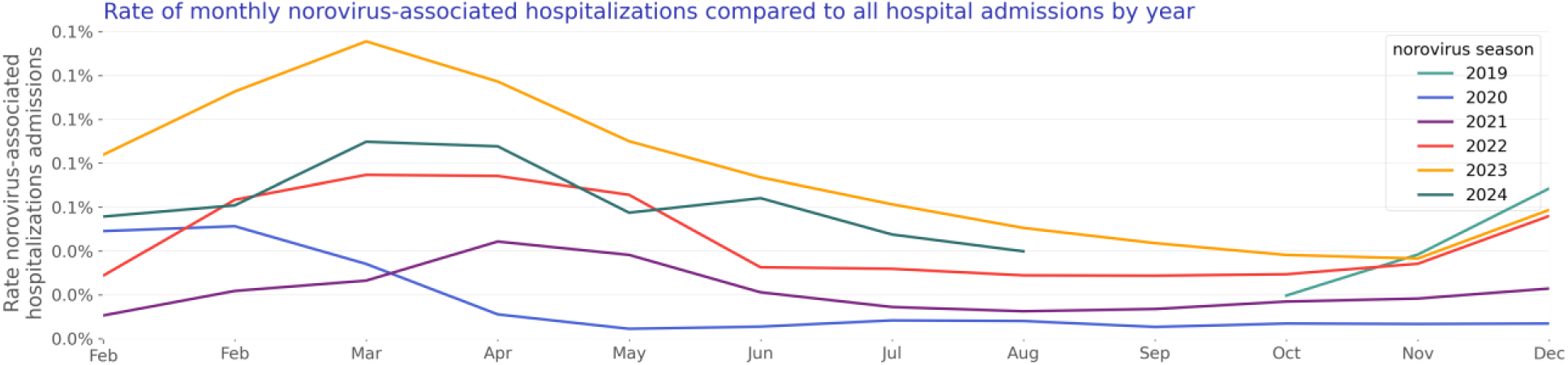
Rate of monthly norovirus-associated hospitalizations compared to all hospital admissions by year

### Test positivity rate over time

We included 166,162 norovirus lab results with known results of which 11,590 were positive. The norovirus test positivity rate is shown in Figure 7. Figure 8 shows yearly trends.

**Figure 7:**
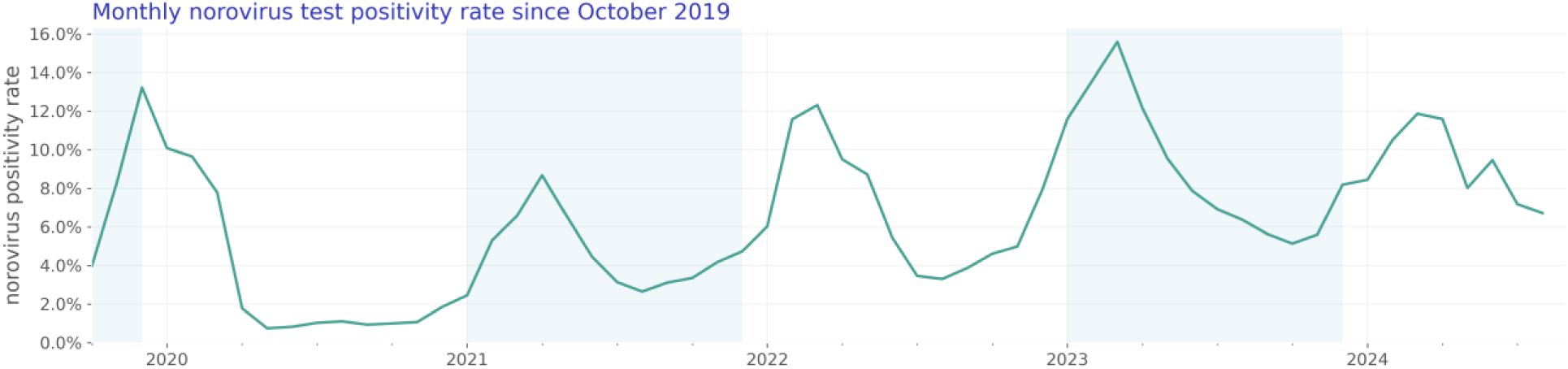
Monthly norovirus test positivity rate since October 2019

**Figure 9:**
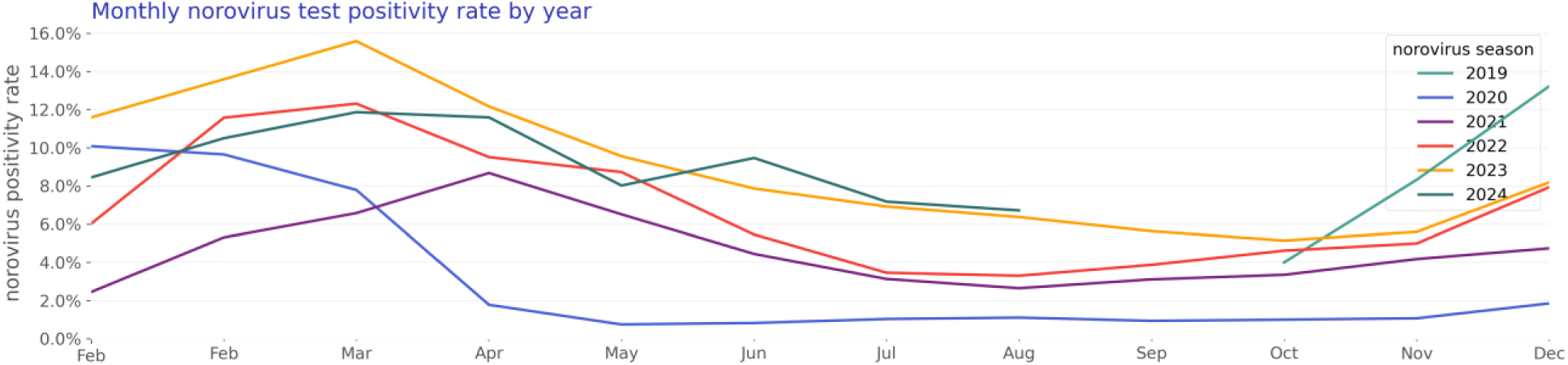
Monthly norovirus test positivity rate by year

### Rotavirus

Our rotavirus study population consists of 1,785 hospitalizations of 1,681 unique patients (Table 3).

**Table 3:**
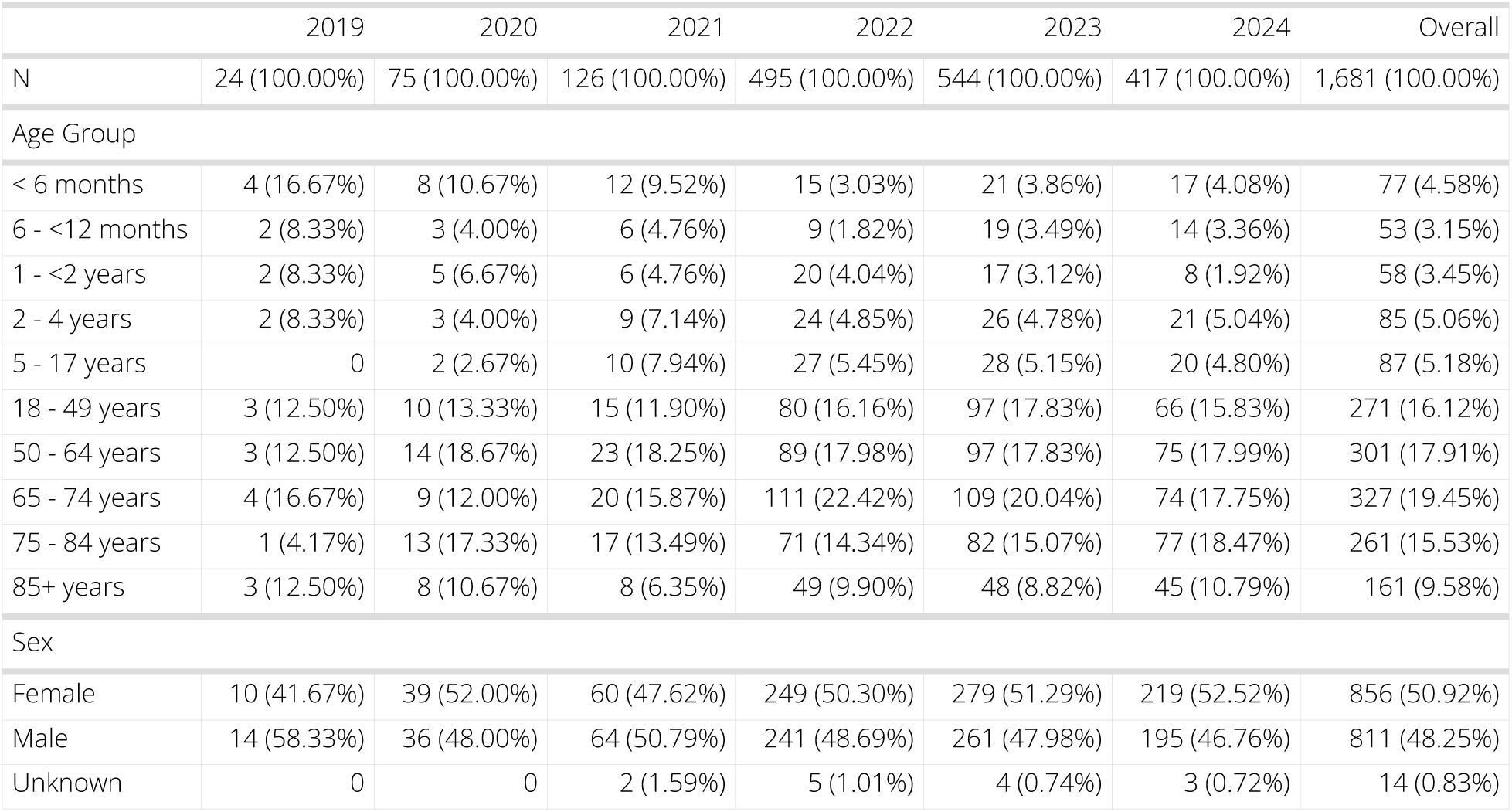
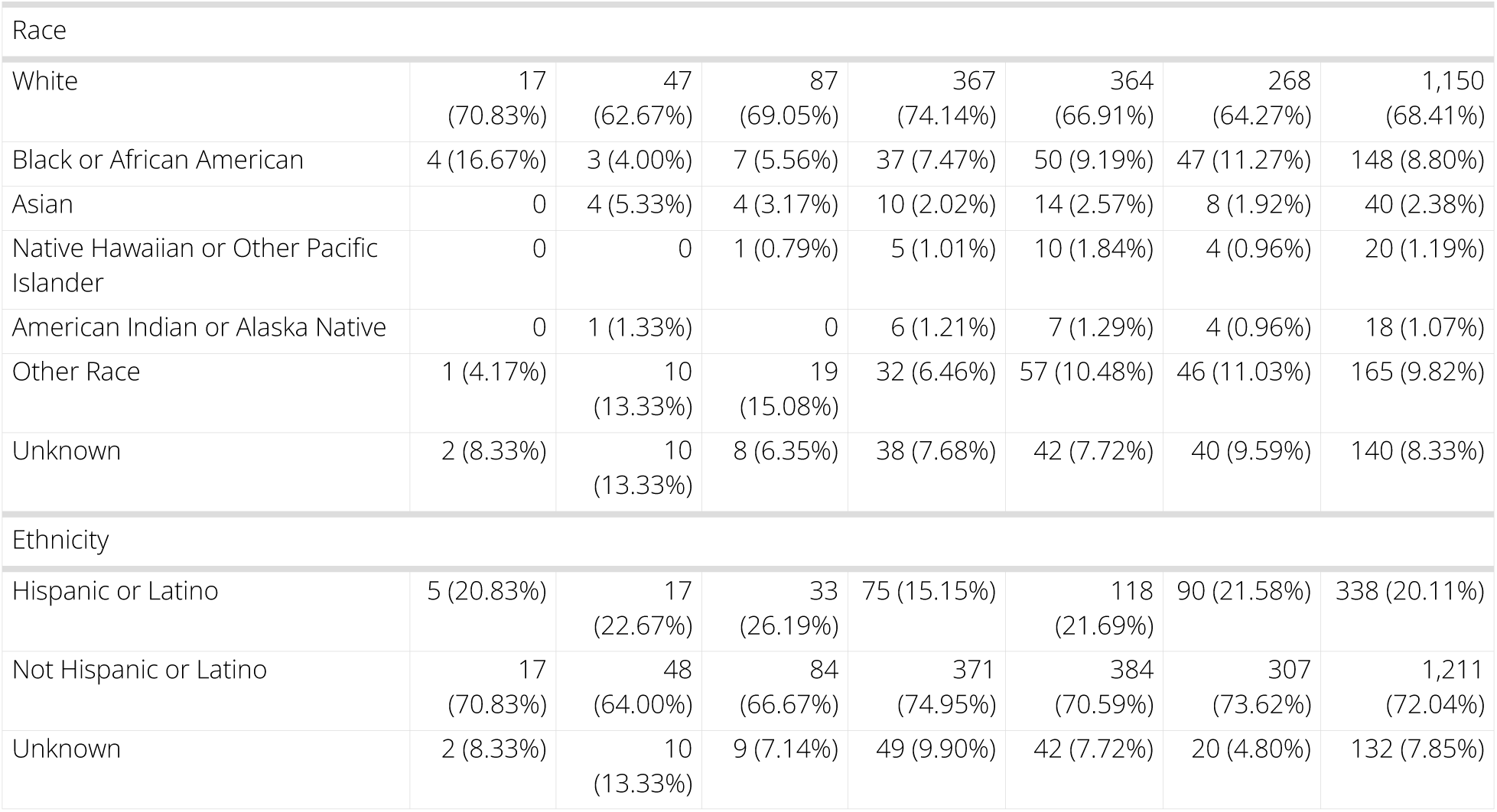
Rotavirus patient characteristics by year.

|  | 2019 | 2020 | 2021 | 2022 | 2023 | 2024 | Overall |
| --- | --- | --- | --- | --- | --- | --- | --- |
| N | 24 (100.00%) | 75 (100.00%) | 126 (100.00%) | 495 (100.00%) | 544 (100.00%) | 417 (100.00%) | 1,681 (100.00%) |
| Age Group |  |  |  |  |  |  |  |
| < 6 months | 4 (16.67%) | 8 (10.67%) | 12 (9.52%) | 15 (3.03%) | 21 (3.86%) | 17 (4.08%) | 77 (4.58%) |
| 6 - <12 months | 2 (8.33%) | 3 (4.00%) | 6 (4.76%) | 9 (1.82%) | 19 (3.49%) | 14 (3.36%) | 53 (3.15%) |
| 1 - <2 years | 2 (8.33%) | 5 (6.67%) | 6 (4.76%) | 20 (4.04%) | 17 (3.12%) | 8 (1.92%) | 58 (3.45%) |
| 2 - 4 years | 2 (8.33%) | 3 (4.00%) | 9 (7.14%) | 24 (4.85%) | 26 (4.78%) | 21 (5.04%) | 85 (5.06%) |
| 5 - 17 years | 0 | 2 (2.67%) | 10 (7.94%) | 27 (5.45%) | 28 (5.15%) | 20 (4.80%) | 87 (5.18%) |
| 18 - 49 years | 3 (12.50%) | 10 (13.33%) | 15 (11.90%) | 80 (16.16%) | 97 (17.83%) | 66 (15.83%) | 271 (16.12%) |
| 50 - 64 years | 3 (12.50%) | 14 (18.67%) | 23 (18.25%) | 89 (17.98%) | 97 (17.83%) | 75 (17.99%) | 301 (17.91%) |
| 65 - 74 years | 4 (16.67%) | 9 (12.00%) | 20 (15.87%) | 111 (22.42%) | 109 (20.04%) | 74 (17.75%) | 327 (19.45%) |
| 75 - 84 years | 1 (4.17%) | 13 (17.33%) | 17 (13.49%) | 71 (14.34%) | 82 (15.07%) | 77 (18.47%) | 261 (15.53%) |
| 85+ years | 3 (12.50%) | 8 (10.67%) | 8 (6.35%) | 49 (9.90%) | 48 (8.82%) | 45 (10.79%) | 161 (9.58%) |
| Sex |  |  |  |  |  |  |  |
| Female | 10 (41.67%) | 39 (52.00%) | 60 (47.62%) | 249 (50.30%) | 279 (51.29%) | 219 (52.52%) | 856 (50.92%) |
| Male | 14 (58.33%) | 36 (48.00%) | 64 (50.79%) | 241 (48.69%) | 261 (47.98%) | 195 (46.76%) | 811 (48.25%) |
| Unknown | 0 | 0 | 2 (1.59%) | 5 (1.01%) | 4 (0.74%) | 3 (0.72%) | 14 (0.83%) |

| Race |  |  |  |  |  |  |  |
| --- | --- | --- | --- | --- | --- | --- | --- |
| White | 17<br>(70.83%) | 47<br>(62.67%) | 87<br>(69.05%) | 367<br>(74.14%) | 364<br>(66.91%) | 268<br>(64.27%) | 1,150<br>(68.41%) |
| Black or African American | 4 (16.67%) | 3 (4.00%) | 7 (5.56%) | 37 (7.47%) | 50 (9.19%) | 47 (11.27%) | 148 (8.80%) |
| Asian | 0 | 4 (5.33%) | 4 (3.17%) | 10 (2.02%) | 14 (2.57%) | 8 (1.92%) | 40 (2.38%) |
| Native Hawaiian or Other Pacific Islander | 0 | 0 | 1 (0.79%) | 5 (1.01%) | 10 (1.84%) | 4 (0.96%) | 20 (1.19%) |
| American Indian or Alaska Native | 0 | 1 (1.33%) | 0 | 6 (1.21%) | 7 (1.29%) | 4 (0.96%) | 18 (1.07%) |
| Other Race | 1 (4.17%) | 10<br>(13.33%) | 19<br>(15.08%) | 32 (6.46%) | 57 (10.48%) | 46 (11.03%) | 165 (9.82%) |
| Unknown | 2 (8.33%) | 10<br>(13.33%) | 8 (6.35%) | 38 (7.68%) | 42 (7.72%) | 40 (9.59%) | 140 (8.33%) |
| Ethnicity |  |  |  |  |  |  |  |
| Hispanic or Latino | 5 (20.83%) | 17<br>(22.67%) | 33<br>(26.19%) | 75 (15.15%) | 118<br>(21.69%) | 90 (21.58%) | 338 (20.11%) |
| Not Hispanic or Latino | 17<br>(70.83%) | 48<br>(64.00%) | 84<br>(66.67%) | 371<br>(74.95%) | 384<br>(70.59%) | 307<br>(73.62%) | 1,211<br>(72.04%) |
| Unknown | 2 (8.33%) | 10<br>(13.33%) | 9 (7.14%) | 49 (9.90%) | 42 (7.72%) | 20 (4.80%) | 132 (7.85%) |

### Hospitalization rate over time

The rate of rotavirus-associated hospitalization is shown in Figure 10. Figure 11 shows yearly trends.

**Figure 10:**
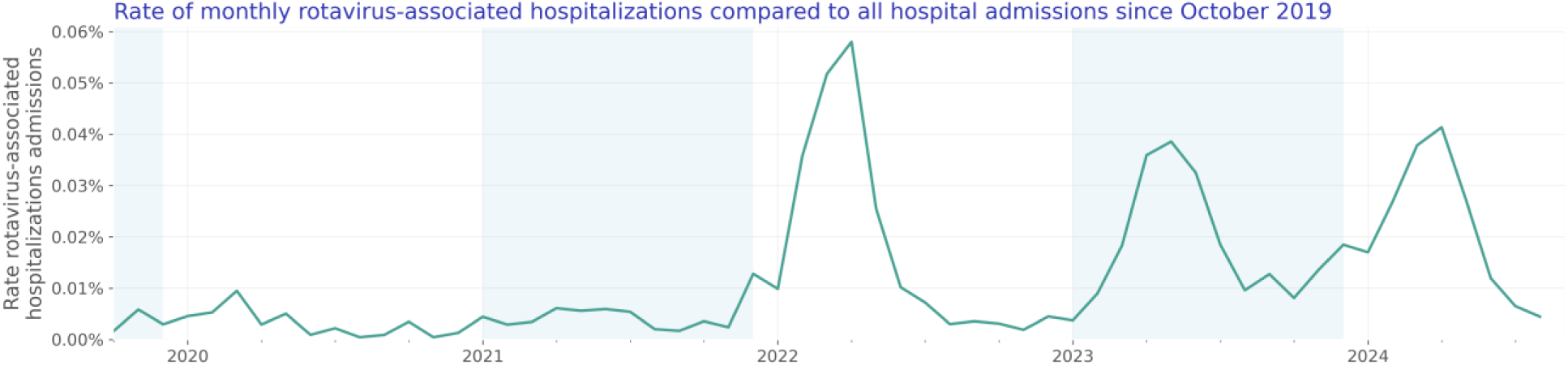
Rate of monthly rotavirus-associated hospitalizations compared to all hospital admissions since October 2019

**Figure 12:**
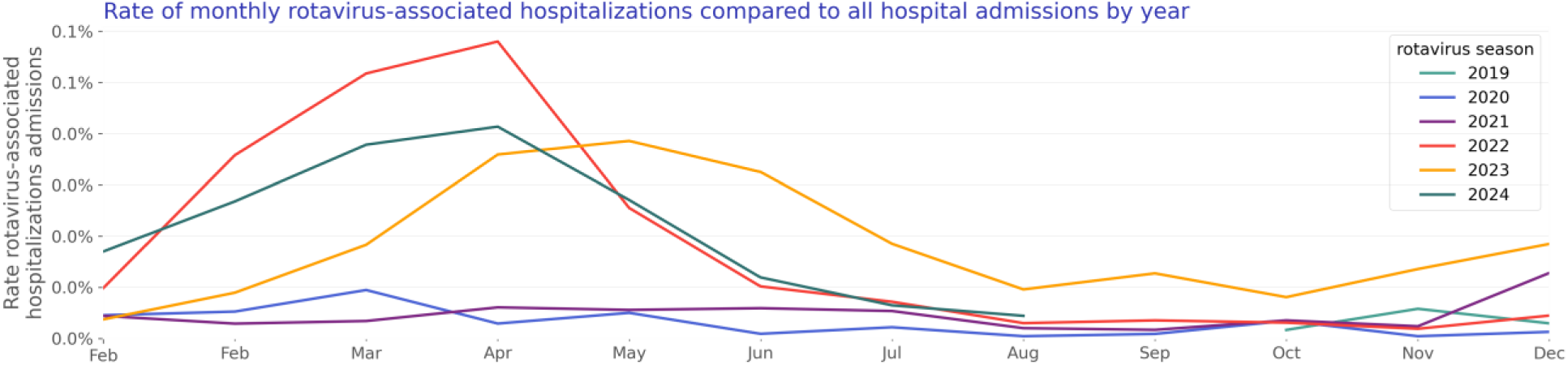
Rate of monthly rotavirus-associated hospitalizations compared to all hospital admissions by year

### Test positivity rate over time

We included 188,073 rotavirus lab results with known results of which 4,151 were positive. The rotavirus test positivity rate is shown in Figure 13. Figure 14 shows yearly trends.

**Figure 13:**
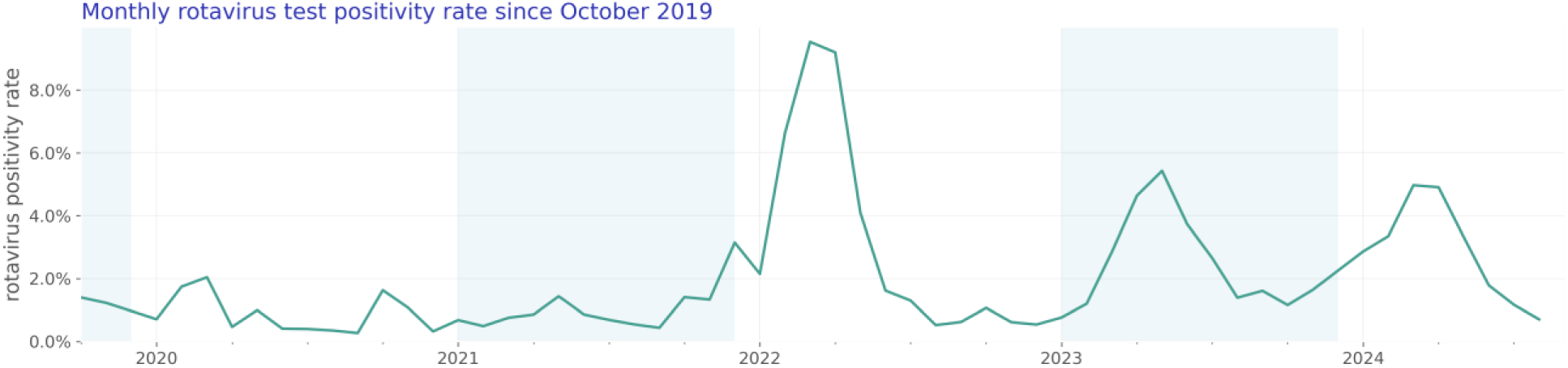
Monthly rotavirus test positivity rate since October 2019

**Figure 15:**
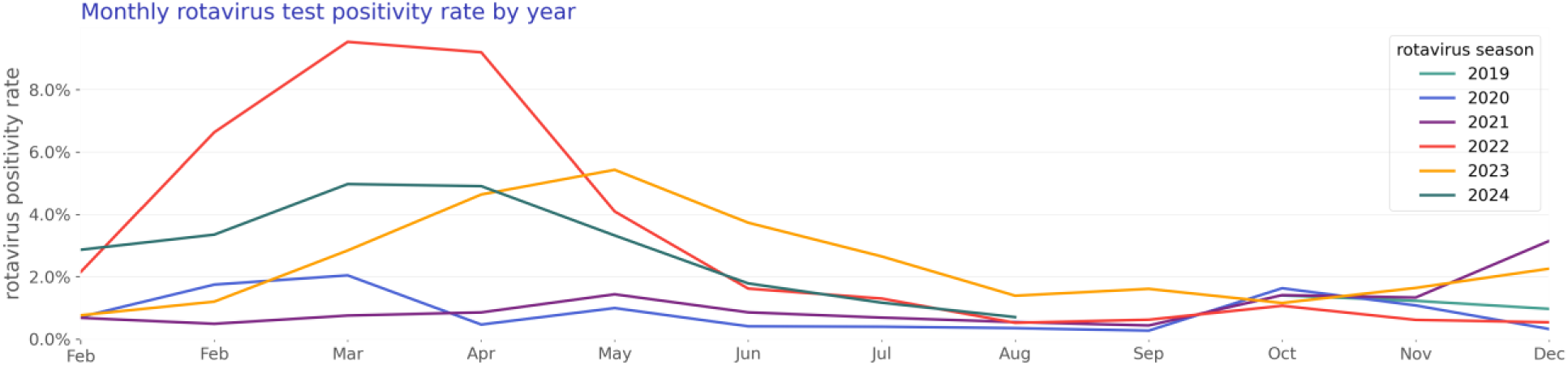
Monthly rotavirus test positivity rate by year

### Infants and children (age 0-4)

Our infants and children study population consists of 692 hospitalizations of 649 unique patients (Table 4).

**Table 4:** Infants and children patient characteristics by year.

|  | 2019 | 2020 | 2021 | 2022 | 2023 | 2024 | Overall |
| --- | --- | --- | --- | --- | --- | --- | --- |
| N | 23<br>(100.00%) | 32<br>(100.00%) | 103<br>(100.00%) | 172<br>(100.00%) | 208<br>(100.00%) | 111<br>(100.00%) | 649<br>(100.00%) |
| Age Group |  |  |  |  |  |  |  |
| < 6 months | 8 (34.78%) | 11 (34.38%) | 24 (23.30%) | 39 (22.67%) | 35 (16.83%) | 23 (20.72%) | 140<br>(21.57%) |
| 6 - <12 months | 3 (13.04%) | 6 (18.75%) | 32 (31.07%) | 30 (17.44%) | 43 (20.67%) | 28 (25.23%) | 142<br>(21.88%) |
| 1 - <2 years | 6 (26.09%) | 8 (25.00%) | 29 (28.16%) | 49 (28.49%) | 56 (26.92%) | 25 (22.52%) | 173<br>(26.66%) |
| 2 - 4 years | 6 (26.09%) | 7 (21.88%) | 18 (17.48%) | 54 (31.40%) | 74 (35.58%) | 35 (31.53%) | 194<br>(29.89%) |
| Sex |  |  |  |  |  |  |  |
| Female | 7 (30.43%) | 9 (28.12%) | 43 (41.75%) | 68 (39.53%) | 92 (44.23%) | 46 (41.44%) | 265<br>(40.83%) |
| Male | 16 (69.57%) | 23 (71.88%) | 59 (57.28%) | 104<br>(60.47%) | 116<br>(55.77%) | 65 (58.56%) | 383<br>(59.01%) |
| Unknown | 0 | 0 | 1 (0.97%) | 0 | 0 | 0 | 1 (0.15%) |
| Race |  |  |  |  |  |  |  |
| White | 15 (65.22%) | 13 (40.62%) | 46 (44.66%) | 86 (50.00%) | 118<br>(56.73%) | 50 (45.05%) | 328<br>(50.54%) |
| Black or African American | 4 (17.39%) | 3 (9.38%) | 14 (13.59%) | 27 (15.70%) | 26 (12.50%) | 13 (11.71%) | 87 (13.41%) |
| Asian | 0 | 3 (9.38%) | 5 (4.85%) | 7 (4.07%) | 13 (6.25%) | 3 (2.70%) | 31 (4.78%) |
| Native Hawaiian or Other<br>Pacific Islander | 0 | 1 (3.12%) | 1 (0.97%) | 10 (5.81%) | 8 (3.85%) | 3 (2.70%) | 23 (3.54%) |
| American Indian or Alaska<br>Native | 0 | 1 (3.12%) | 1 (0.97%) | 0 | 2 (0.96%) | 1 (0.90%) | 5 (0.77%) |
| Other Race | 2 (8.70%) | 7 (21.88%) | 29 (28.16%) | 27 (15.70%) | 26 (12.50%) | 23 (20.72%) | 114<br>(17.57%) |
| Unknown | 2 (8.70%) | 4 (12.50%) | 7 (6.80%) | 15 (8.72%) | 15 (7.21%) | 18 (16.22%) | 61 (9.40%) |
| Ethnicity |  |  |  |  |  |  |  |
| Hispanic or Latino | 8 (34.78%) | 12 (37.50%) | 35 (33.98%) | 46 (26.74%) | 80 (38.46%) | 45 (40.54%) | 226<br>(34.82%) |
| Not Hispanic or Latino | 15 (65.22%) | 17 (53.12%) | 63 (61.17%) | 113<br>(65.70%) | 117<br>(56.25%) | 56 (50.45%) | 381<br>(58.71%) |
| Unknown | 0 | 3 (9.38%) | 5 (4.85%) | 13 (7.56%) | 11 (5.29%) | 10 (9.01%) | 42 (6.47%) |
| Virus |  |  |  |  |  |  |  |
| norovirus | 13 (56.52%) | 13 (40.62%) | 70 (67.96%) | 104<br>(60.47%) | 125<br>(60.10%) | 51 (45.95%) | 376<br>(57.94%) |
| rotavirus | 10 (43.48%) | 19 (59.38%) | 33 (32.04%) | 68 (39.53%) | 83 (39.90%) | 60 (54.05%) | 273<br>(42.06%) |

### Hospitalization rate over time

The rate of viral gastroenteritis-associated hospitalizations compared to all hospitalizations for infants and children under age five is shown in Figure 16. Figure 17 shows the same data stacked to represent the combined impact of the viruses.

**Figure 16:**
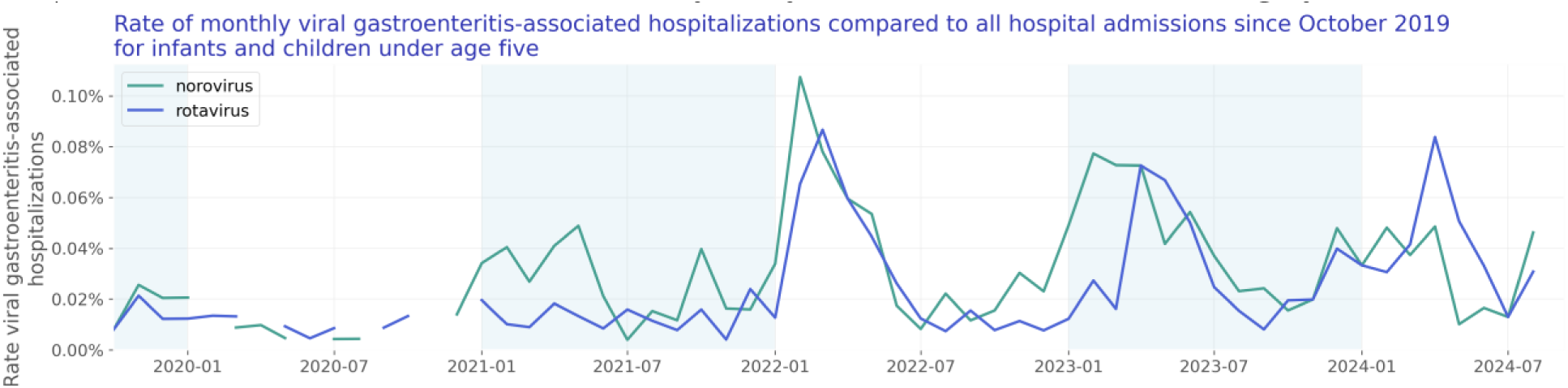
Rate of monthly viral gastroenteritis-associated hospitalizations compared to all hospital admissions since October 2019 for infants and children under age five

**Figure 17:**
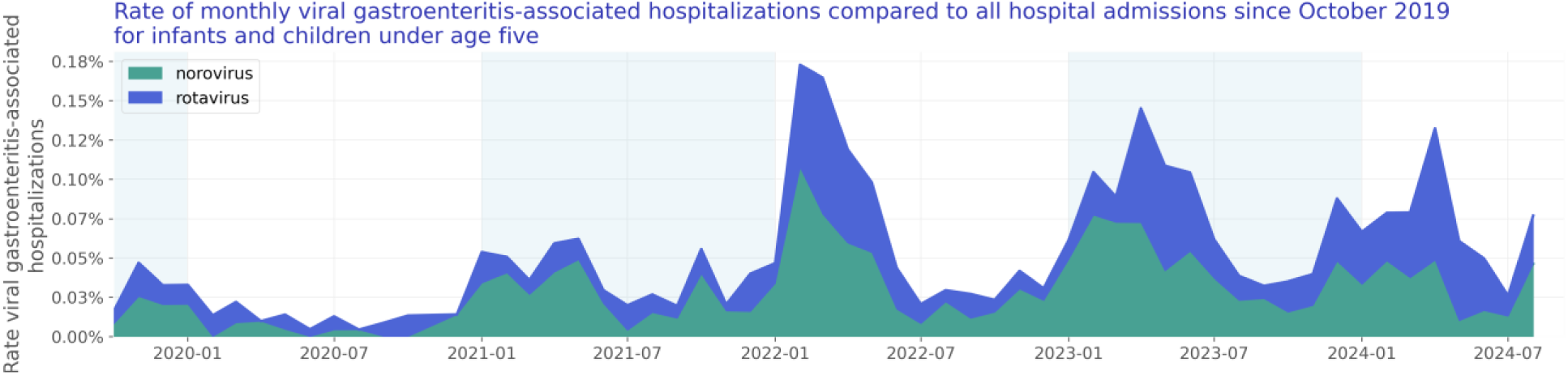
Rate of monthly viral gastroenteritis-associated hospitalizations compared to all hospital admissions since October 2019 for infants and children under age five

### Test positivity rate over time

We included 16,838 lab results with known results of infants and children under age five. Of those tests, 2,177 lab results were positive. The test positivity rate for infants and children under age five is shown in Figure 18.

**Figure 18:**
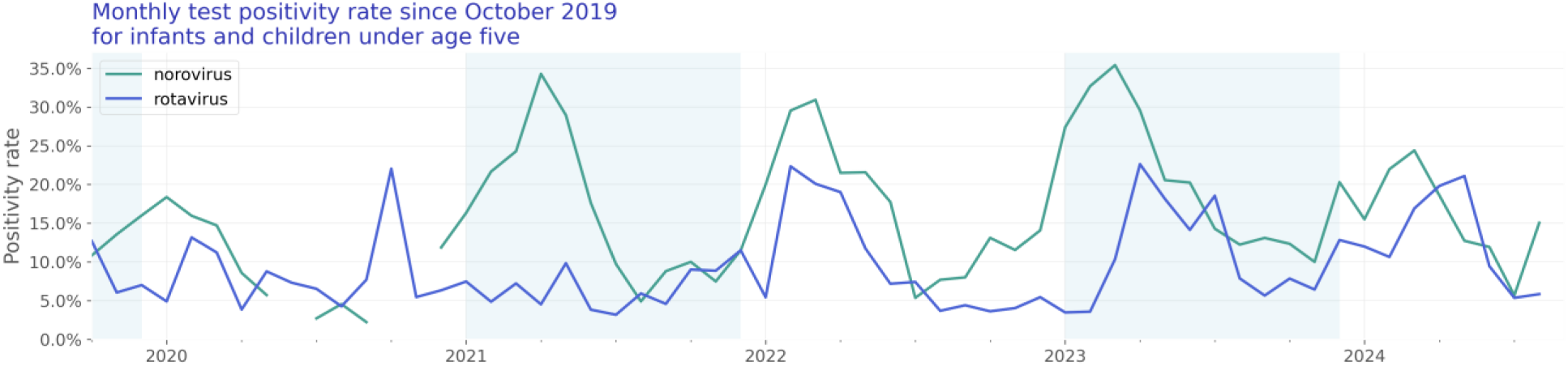
Monthly test positivity rate since October 2019 for infants and children under age five

### Older adults (aged 65 and over)

Our older adults study population consists of 3,922 hospitalizations of 3,841 unique patients (Table 5).

**Table 5:** Older adults patient characteristics by year.

|  | 2019 | 2020 | 2021 | 2022 | 2023 | 2024 | Overall |
| --- | --- | --- | --- | --- | --- | --- | --- |
| N | 138<br>(100.00%) | 243<br>(100.00%) | 289<br>(100.00%) | 866 (100.00%) | 1,457<br>(100.00%) | 848<br>(100.00%) | 3,841<br>(100.00%) |
| Age Group |  |  |  |  |  |  |  |
| 65 - 74 years | 51 (36.96%) | 81 (33.33%) | 109<br>(37.72%) | 380 (43.88%) | 592<br>(40.63%) | 322<br>(37.97%) | 1,535<br>(39.96%) |
| 75 - 84 years | 53 (38.41%) | 105<br>(43.21%) | 120<br>(41.52%) | 297 (34.30%) | 524<br>(35.96%) | 332<br>(39.15%) | 1,431<br>(37.26%) |
| 85+ years | 34 (24.64%) | 57 (23.46%) | 60 (20.76%) | 189 (21.82%) | 341<br>(23.40%) | 194<br>(22.88%) | 875<br>(22.78%) |
| Sex |  |  |  |  |  |  |  |
| Female | 74 (53.62%) | 141<br>(58.02%) | 173<br>(59.86%) | 494 (57.04%) | 807<br>(55.39%) | 511<br>(60.26%) | 2,200<br>(57.28%) |
| Male | 64 (46.38%) | 101<br>(41.56%) | 115<br>(39.79%) | 368 (42.49%) | 642<br>(44.06%) | 329<br>(38.80%) | 1,619<br>(42.15%) |
| Unknown | 0 | 1 (0.41%) | 1 (0.35%) | 4 (0.46%) | 8 (0.55%) | 8 (0.94%) | 22 (0.57%) |
| Race |  |  |  |  |  |  |  |
| White | 123<br>(89.13%) | 197<br>(81.07%) | 237<br>(82.01%) | 735 (84.87%) | 1,174<br>(80.58%) | 653<br>(77.00%) | 3,119<br>(81.20%) |
| Black or African American | 6 (4.35%) | 17 (7.00%) | 20 (6.92%) | 43 (4.97%) | 74 (5.08%) | 64 (7.55%) | 224 (5.83%) |
| Asian | 0 | 1 (0.41%) | 2 (0.69%) | 7 (0.81%) | 28 (1.92%) | 11 (1.30%) | 49 (1.28%) |
| Native Hawaiian or Other Pacific Islander | 0 | 0 | 0 | 0 | 3 (0.21%) | 2 (0.24%) | 5 (0.13%) |
| American Indian or Alaska Native | 0 | 1 (0.41%) | 2 (0.69%) | 4 (0.46%) | 9 (0.62%) | 5 (0.59%) | 21 (0.55%) |
| Other Race | 3 (2.17%) | 6 (2.47%) | 9 (3.11%) | 24 (2.77%) | 55 (3.77%) | 43 (5.07%) | 140 (3.64%) |
| Unknown | 6 (4.35%) | 21 (8.64%) | 19 (6.57%) | 53 (6.12%) | 114 (7.82%) | 70 (8.25%) | 283 (7.37%) |
| Ethnicity |  |  |  |  |  |  |  |
| Hispanic or Latino | 2 (1.45%) | 9 (3.70%) | 18 (6.23%) | 47 (5.43%) | 109 (7.48%) | 70 (8.25%) | 255 (6.64%) |
| Not Hispanic or Latino | 126<br>(91.30%) | 203<br>(83.54%) | 239<br>(82.70%) | 730<br>(84.30%) | 1,217<br>(83.53%) | 727<br>(85.73%) | 3,242 (84.41%) |
| Unknown | 10 (7.25%) | 31 (12.76%) | 32 (11.07%) | 89<br>(10.28%) | 131 (8.99%) | 51 (6.01%) | 344 (8.96%) |

| Virus |  |  |  |  |  |  |  |
| --- | --- | --- | --- | --- | --- | --- | --- |
| norovirus | 130 (94.20%) | 213 (87.65%) | 244 (84.43%) | 635 (73.33%) | 1,218 (83.60%) | 652 (76.89%) | 3,092 (80.50%) |
| rotavirus | 8 (5.80%) | 30 (12.35%) | 45 (15.57%) | 231 (26.67%) | 239 (16.40%) | 196 (23.11%) | 749 (19.50%) |

### Hospitalization rate over time

The rate of viral gastroenteritis-associated hospitalizations compared to all hospitalizations for adults aged 65 and over is shown in Figure 19. Figure 20 shows the same data stacked to represent the combined impact of the viruses.

**Figure 19:**
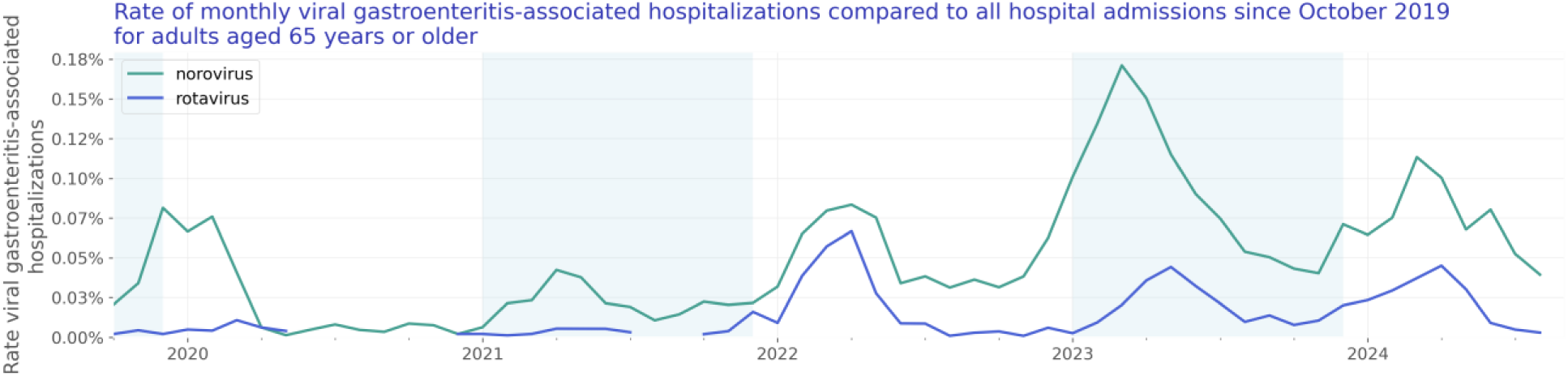
Rate of monthly viral gastroenteritis-associated hospitalizations compared to all hospital admissions since October 2019 for adults aged 65 years or older

**Figure 20:**
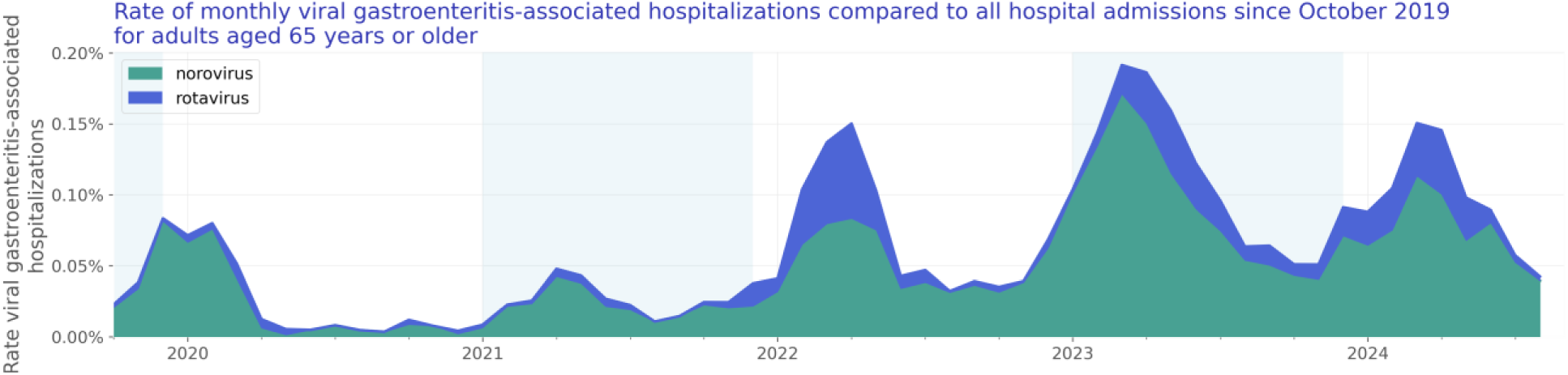
Rate of monthly viral gastroenteritis-associated hospitalizations compared to all hospital admissions since October 2019 for adults aged 65 years or older

### Test positivity rate over time

We included 153,590 lab results with known results of adults aged 65 and over. Of those tests, 5,057 lab results were positive. The test positivity rate for adults aged 65 and over is shown in Figure 21.

**Figure 19:**
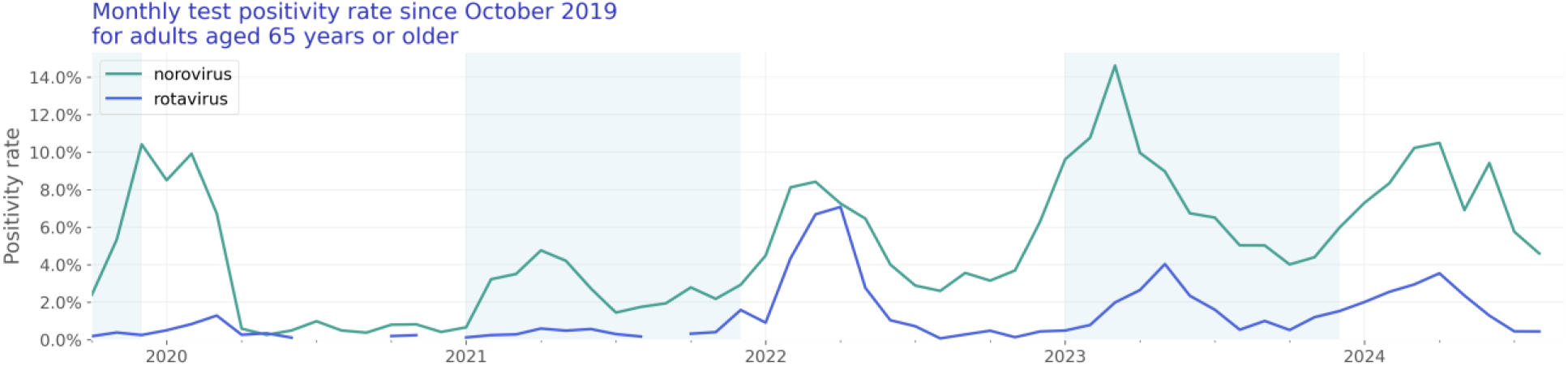
Rate of monthly viral gastroenteritis-associated hospitalizations compared to all hospital admissions since October 2019 for adults aged 65 years or older

## Discussion and Conclusion

We see evidence that we are in the nadir of the norovirus and rotavirus season for the overall population. The overall timeseries trends of hospitalizations associated with these viruses show a steady decrease over the last months, leading to a rate of 0.04% in August 2024 in the overall population. However, we see first indicators that the next season is already beginning for the population age 0-4 years old with an increase of the hospitalization rate of +196.7% from July.

Several limitations of our analysis should be noted. All data are preliminary and may change as additional data are obtained. These findings are consistent with data accessed September 09, 2024. These are raw counts and post-stratification methods have not been conducted. This analysis does not include patients hospitalized with a viral gastroenteritis who were not tested or were tested later in their medical care (when laboratory tests results would have returned a negative result). In addition, cohorts with small counts may be suppressed during the de-identification process leading to the appearance of zero patients for a given time period. Moreover, the unknowns in this report either indicate the value was not included in the individual’s electronic health record or that it was excluded from the data to protect an individual’s identity as a part of Truveta’s commitment to privacy (Truveta, 2024).

We will continue to monitor viral gastroenteritis-associated hospitalization overall and for the at-risk populations.

## Suggested citation

Suggested citation: “Truveta Monitoring Report: Viral gastroenteritis - August 2024 Data, Truveta Inc. Truveta.com/research. Accessed on DATE”.

## Data Availability

The data used in this study are available to all Truveta subscribers and may be accessed at studio.truveta.com

